# Pelvic Stability During Total Hip Arthroplasty: A Comparison of Traction and Standard Surgical Tables Using Augmented Reality Navigation

**DOI:** 10.1101/2024.09.21.24314140

**Authors:** Musashi Ima, Tamon Kabata, Daisuke Inoue, Yu Yanagi, Takahiro Iyobe, Naoya Fujimaru, Satoru Demura

**Author notes:** **Corresponding author:** Tamon Kabata, PhD Department of Orthopedic Surgery, Graduate School of Medical Science, Kanazawa University 13-1 Takaramachi, Kanazawa, Ishikawa 920-8641, Japan.

## Abstract

**Background:** Total hip arthroplasty enhances quality of life by improving joint function. However, the success of this procedure heavily relies on precise acetabular cup positioning, which is insufficient using traditional placement guidelines. This insufficiency is evidenced by the persistent occurrence of dislocations, highlighting the critical need for enhanced pelvic alignment accuracy. Standard surgical tables present challenges in maintaining consistent pelvic positioning during the procedure, making it difficult to achieve optimal alignment.

**Purpose:** This study aimed to evaluate the effectiveness of a traction surgical table in stabilizing pelvic movements during total hip arthroplasty and compare it with the use of a standard surgical table.

**Methods:** This retrospective study assessed 88 total hip arthroplasties performed using the AR-HIP (Zimmer Biomet Japan, Tokyo, Japan) system for real-time pelvic navigation. Procedures were performed using either a standard surgical table (n=48) or a traction surgical table (n=40). Pelvic alignment was monitored at multiple surgical stages, and stability was statistically analyzed to compare the efficacy of the two table types.

**Results:** The traction table significantly stabilized anteroposterior pelvic movements across all critical surgical stages (p<0.05), except during anterior capsular release. In contrast, procedures performed on the standard table exhibited less stability in anteroposterior pelvic movements, suggesting the superior performance of the traction table.

**Conclusions:** Compared with the standard table, the use of the traction table in total hip arthroplasty effectively controls anteroposterior pelvic movements and improves acetabular cup alignment. However, lateral pelvic stability remains uncontrolled using both table types, indicating a need for further technological advancements in surgical practice to improve the outcomes of total hip arthroplasty.

## 1. Introduction

Total hip arthroplasty (THA) substantially enhances a patient’s quality of life by restoring joint function [1,2]. The success of this procedure depends on various factors, with precise acetabular cup positioning being of paramount importance. Proper placement mitigates the risk of dislocations and reduces polyethylene wear; thus, it plays a crucial role in the long-term success of THA [3]. The “safe zone” proposed by Lewinnek et al in 1978, who recommended an inclination of 30–50° and anteversion of 5–25°, serves as a benchmark for acetabular component orientation [4]. Nevertheless, even placement within this zone is associated with dislocations [5], suggesting that the ideal zone may be narrower [6].

The pelvic tilt affects the alignment of the acetabular component. A previous study reported a linear correlation between pelvic angle and cup anteversion, with a 1° change in pelvic tilt altering anteversion by approximately 0.74°, as well as a nonlinear relationship between pelvic tilt and cup inclination, with a 1° change in pelvic angle resulting in an average inclination adjustment of 0.29° [7]. This highlights the importance of understanding pelvic positioning during surgery to anticipate its impact on cup placement.

Recently, the anterior approach in the supine position has gained popularity for THA [8]. This approach preserves muscle integrity, facilitates faster functional recovery, and potentially reduces the length of hospital stay [9]. However, both patient factors (e.g., body mass index [BMI], anatomical characteristics) and surgical factors (e.g., incision size, field of view) can impede a surgeon’s ability to accurately position the acetabular cup [10], and even experienced surgeons may encounter numerous outliers in freehand supine THA [11].

Traditional surgical tables often present challenges in maintaining consistent pelvic positioning during the procedure, which can compromise the accuracy of acetabular cup placement. In contrast, traction tables can more effectively stabilize the pelvis, potentially leading to better surgical outcomes. Furthermore, the AR-HIP system (Zimmer Biomet Japan, Tokyo, Japan) is utilized to prevent acetabular cup misalignment. This portable navigation device enables surgeons to use a smartphone app to align the cup based on recorded landmarks and construct three-dimensional coordinates for precise placement. In this system, the functional pelvic plane (FPP) is defined as a line between the direction of gravity and the anterior superior iliac spine. With the AR-HIP system, surgeons can overlay the FPP onto the actual operative field displayed on a smartphone, allowing for real-time monitoring and adjustment of cup orientation. Pelvic movements can also be observed and assessed throughout the procedure (Figure 1) [12].

**Figure 1.**
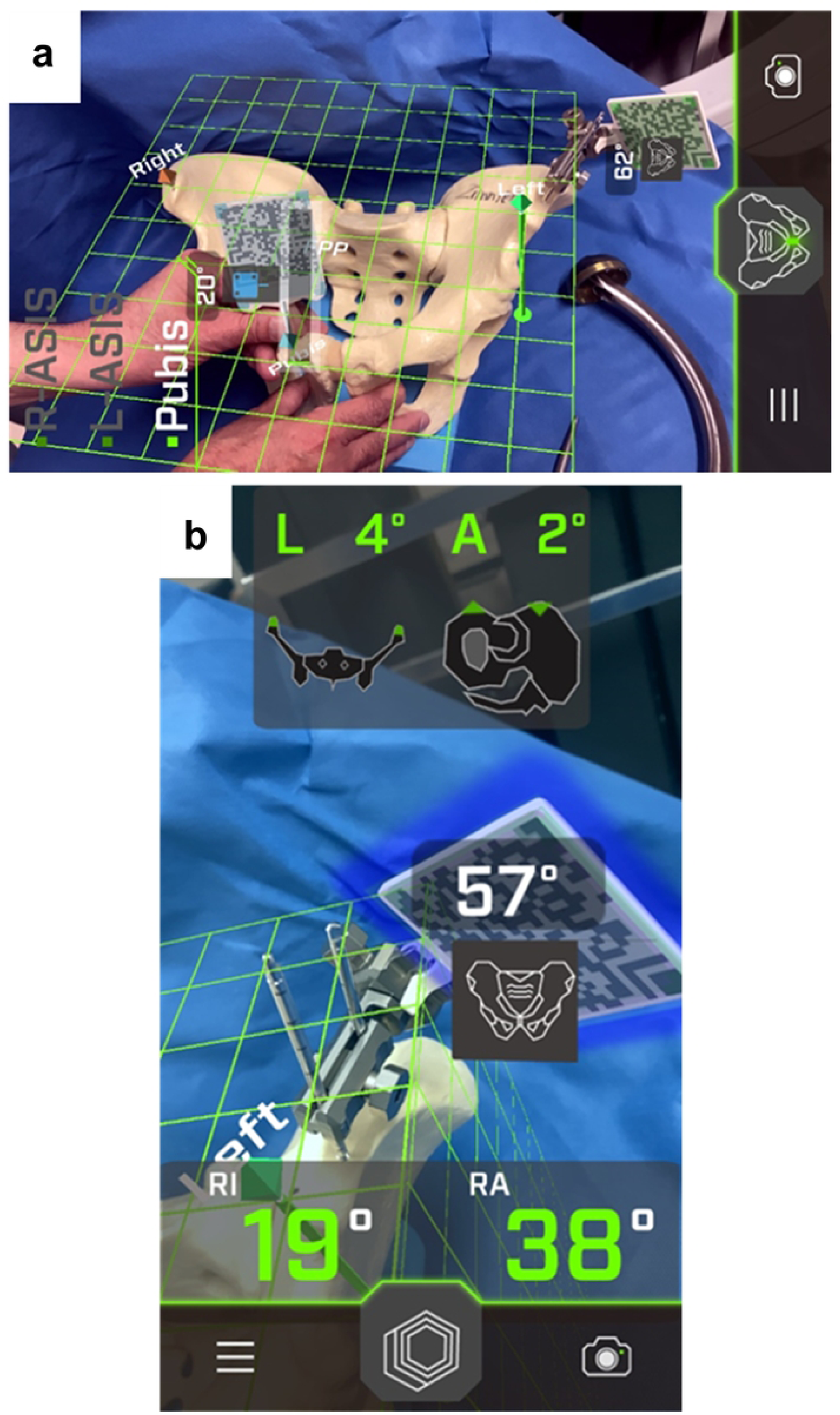
(a) Registration photo: Recognition of the anatomical pelvic plane through landmarks such as the bilateral anterior superior iliac spines and the pubic symphysis. The functional pelvic plane is calculated using the built-in gyro sensor of the smartphone. (b)Intraoperatively: The smartphone mounted on the cup handle scans a QR code placed on the pelvis, displaying real-time data on cup alignment and pelvic tilt in both the anteroposterior and lateral directions.

The current study aimed (1) to observe and quantify pelvic movements at various stages of THA using the AR-HIP system to identify potential shifts in pelvic alignment and (2) to determine the extent of improvement in pelvic alignment when a traction table is used, as compared with conventional surgical tables.

## 2. Materials and methods

### 2.1. Patients

This retrospective single-center study initially included 102 hip joints treated for osteoarthritis (OA) at our institution from December 2022 to February 2024. However, patients with inflammatory diseases or severe deformities (Crowe types 3 and 4) and those who underwent revision THA were excluded, resulting in 88 eligible hip joints. Patient records were retrospectively reviewed to collect demographic data (age, sex, and BMI) and other variables (operative time, perioperative and postoperative complications, and length of hospital stay). Complications were defined as intraoperative fractures, postoperative fractures or subsidence, dislocations, wound complications, infections, abscesses, nerve damage, or the need for reoperation. The data were anonymized and stored in a secure electronic database.

### 2.2. Surgical procedure

All procedures were performed by a single surgeon with 20 years of experience in orthopedic surgery. The anterolateral approach was used, and the implant was selected based on the surgeon’s preference without alteration from standard procedures. No randomization or selection of patients was performed. A total of 48 and 40 procedures were performed on a standard operating table from December 2022 to March 2023 and on a traction table (RotexTable®) from September 2023 to March 2024, respectively. Allocation to the standard or traction table was based on the operating room’s availability and the surgeon’s preference. No specific patient characteristics determined the choice of table, thereby minimizing potential selection bias. The cup was placed to replicate the center of the contralateral femoral head and was internalized until it contacted the teardrop. The stem was positioned to extend the offset corresponding to the internalization of the cup, aligning the global offset with the contralateral side.

### 2.3. Pelvic alignment measurement using the AR-HIP system

The surgeon used the AR-HIP system, a portable navigation device, through a smartphone app (AR-HIP; Zimmer Biomet, Tokyo, Japan). Specific landmarks were recorded to establish three-dimensional coordinates for precise cup positioning. The FPP was defined as a line between the direction of gravity and the anterior superior iliac spine. This allowed for real-time visualization and adjustment of cup orientation, with pelvic movements monitored and assessed throughout the procedure. The baseline for pelvic alignment was set as the FPP at preoperative registration, with subsequent measurements taken after anterior capsular release, femoral head resection, cup placement, and post-reduction once the retractors were removed.

Details on the applied traction force, limb positioning on the traction table, and the specific components of pelvic rotational movement measured were meticulously documented. The study focused on rotations in the sagittal and transverse planes (anteroposterior and lateral directions, respectively), whereas coronal plane (vertical axis) rotations were not evaluated.

### 2.4. Data analysis

Statistical analyses included an independent sample t-test for continuous variables and the chi-square test for categorical variables. Non-parametric tests (Mann–Whitney U test and Fisher’s exact test) were conducted as needed. Normality was assessed using the Shapiro– Wilk test. Potential confounding variables such as age, BMI, and preoperative joint function were evaluated and controlled for in the analysis to ensure that they did not significantly affect the results. Statistical significance was set at p<0.05. All tests were two-tailed. Data were analyzed using SPSS version 28 (IBM Corp., Armonk, NY, USA).

### 2.5. Ethical Considerations

This study adhered to the principles of the Declaration of Helsinki. The need for ethics approval was waived by the Medical Ethics Committee of Kanazawa University according to the “Ethical Guidelines for Medical and Health Research Involving Human Subjects” provided by the Ministry of Education, Culture, Sports, Science and Technology (MEXT) and the Ministry of Health, Labour and Welfare (MHLW) in Japan. Written informed consent was obtained from all participants. The data were accessed on 20^th^ February, 2024 for research purposes. The authors did not have access to information that could identify individual participants during or after data collection. Confidentiality and anonymity were ensured, and participants were informed of their right to withdraw at any time.

## 3. Results

### 3.1. Patient demographics

The demographic characteristics of the patients are summarized in Table 1. Overall, the standard table group comprised 48 patients, whereas the traction table group consisted of 40 patients. The distributions of osteoarthritis and osteonecrosis of the femoral head were similar between the groups, with 40 osteoarthritis cases and 8 osteonecrosis of the femoral head cases in the standard table group and 36 osteoarthritis cases and 4 osteonecrosis of the femoral head cases in the traction table group (p=0.36).

**Table 1.**
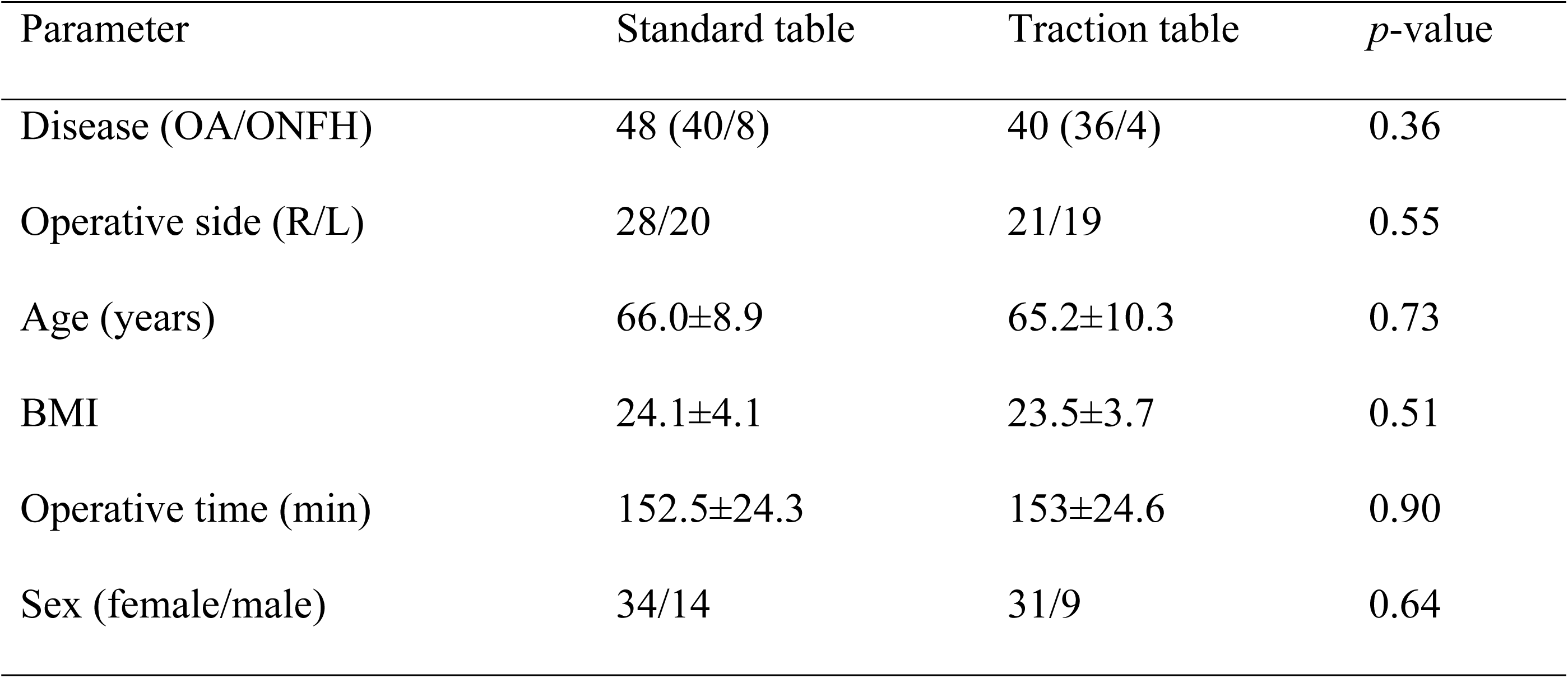
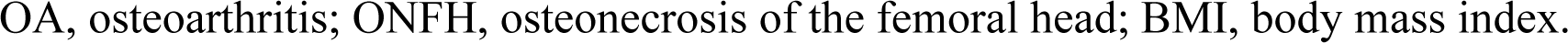
Demographic data of patients.

| Parameter | Standard table | Traction table | *p*-value |
| --- | --- | --- | --- |
| Disease (OA/ONFH) | 48 (40/8) | 40 (36/4) | 0.36 |
| Operative side (R/L) | 28/20 | 21/19 | 0.55 |
| Age (years) | 66.0±8.9 | 65.2±10.3 | 0.73 |
| BMI | 24.1±4.1 | 23.5±3.7 | 0.51 |
| Operative time (min) | 152.5±24.3 | 153±24.6 | 0.90 |
| Sex (female/male) | 34/14 | 31/9 | 0.64 |

OA, osteoarthritis; ONFH, osteonecrosis of the femoral head; BMI, body mass index.

The two groups showed no significant difference with respect to the operative side (right/left), with 28 right hips and 20 left hips in the standard table group and 21 right hips and 19 left hips in the traction table group (p=0.55). The average patient age was 66.0±8.9 years and 65.2±10.3 years in the standard and traction table groups, respectively (p=0.73). BMI was 24.1±4.1 kg/m^2^ and 23.5±3.7 kg/m^2^ in the standard and traction table groups, respectively (p=0.51). The average operative time was 152.5±24.3 min and 153±24.6 min in the standard and traction table groups, respectively (p=0.90). The sex distribution was also similar, with 34 females and 14 males in the standard table group and 31 females and 9 males in the traction table group (p=0.64).

### 3.2. Intraoperative pelvic alignment changes

#### 3.2.1. Without a traction table

The changes in pelvic alignment in the anteroposterior direction during THA without the use of a traction table are shown in Figure 2. In THA, the pelvis tilted forward by 0.9±1.3° on average during anterior capsular release, 3.4±0.8° during femoral head resection, 2.3±0.8° during cup placement, and 2.0±0.8° during reduction. The change averaged 0.9±1.3° from AR registration to anterior capsular release, 2.52±1.4° from anterior capsular release to femoral head resection, -1.1±0.9° from femoral head resection to cup placement, and - 0.3±0.7° from cup placement to reduction. The greatest forward tilt occurred between the anterior capsular release and femoral head resection (p<0.05).

**Figure 2.**
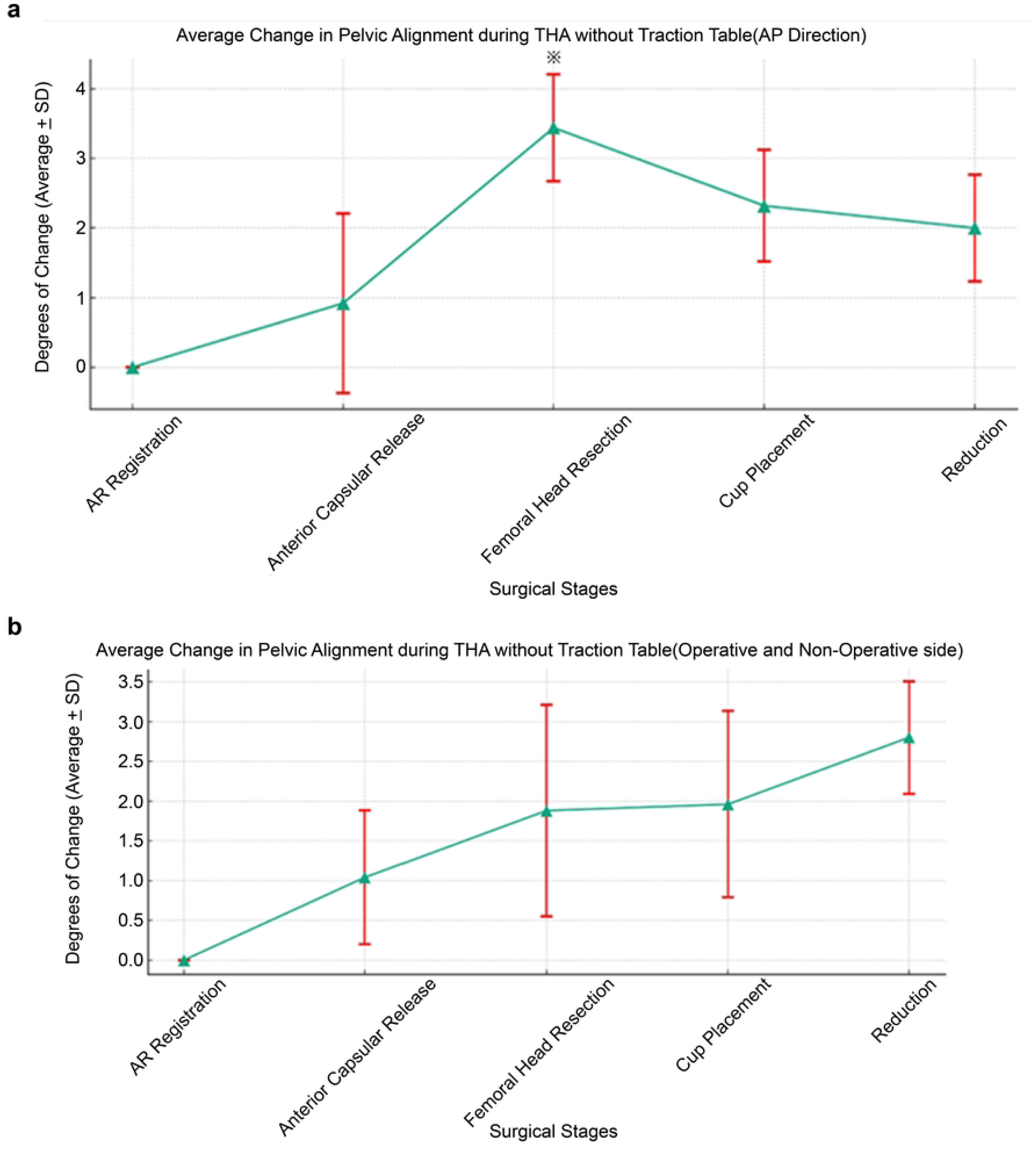
(a) Pelvic tilt increased anteriorly from registration through each surgical stage, with the most significant change occurring between anterior capsular release and femoral head resection (mean change: 2.52±1.4°, p<0.05). (b) Lateral tilt towards the operative side also showed significant variation, particularly after cup placement (p=0.029).

The changes in lateral pelvic alignment (operative and non-operative sides) intraoperatively are shown in Figure 3. The pelvis tilted laterally towards the operative side by 1.0±0.8° on average during anterior capsular release, 1.9±1.3° during femoral head resection, 2.0±1.3° during cup placement, and 2.8±0.7° during reduction. Throughout the procedure, the pelvis tended to tilt towards the operative side. The change averaged 1.0±0.8° from AR registration to anterior capsular release, 0.8±1.3° from anterior capsular release to femoral head resection, 0.4±1.58° from femoral head resection to cup placement, and 0.8±0.9° from cup placement to reduction. In general, the changes in lateral pelvic movement were minimal. However, the variability observed during cup placement and reduction suggests a greater degree of instability in the lateral direction, which could lead to potential malalignment in the absence of adequate control (Figure 2).

**Figure 3.**
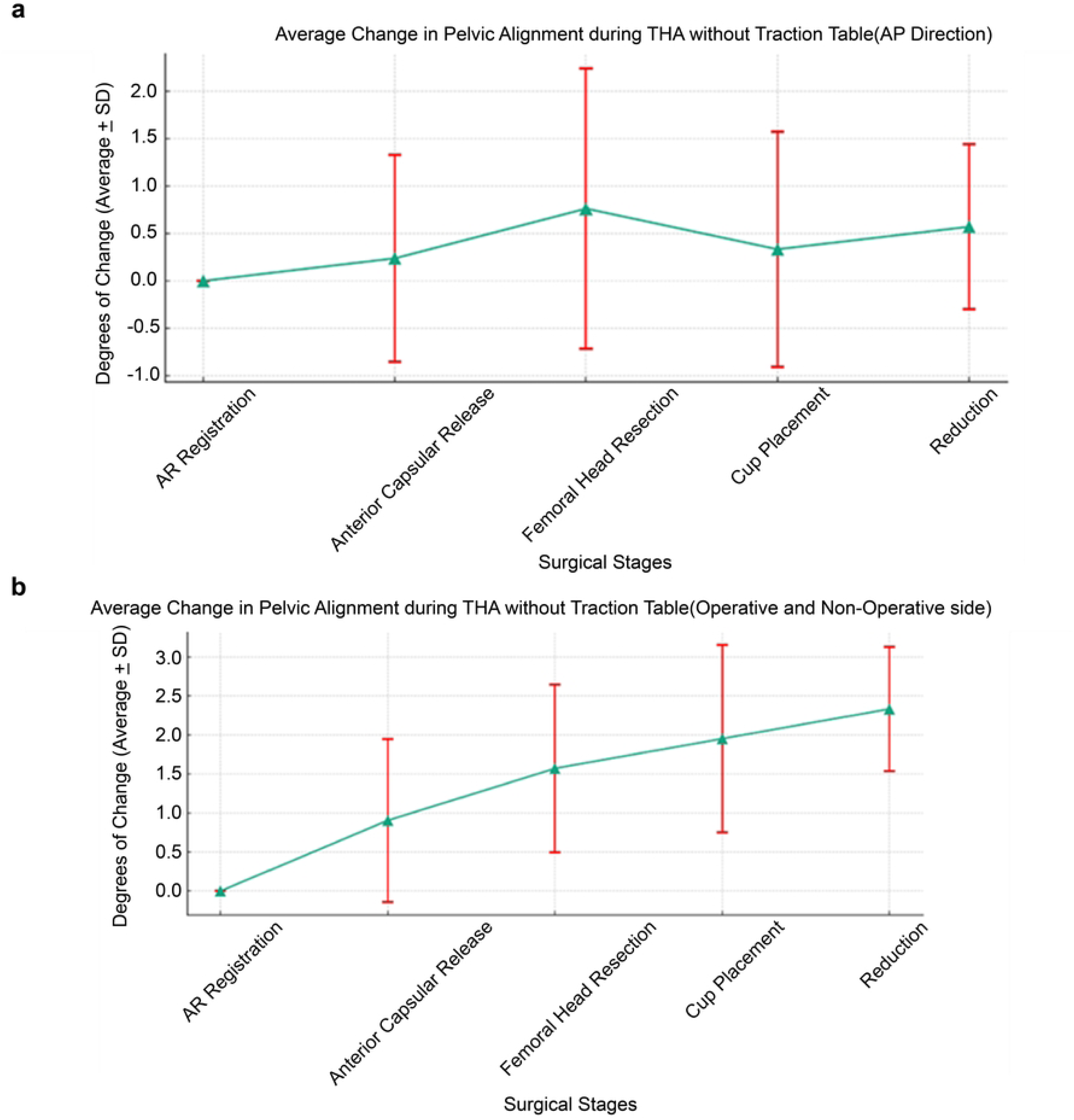
(a) The use of a traction table significantly stabilized the anteroposterior pelvic tilt across all stages of surgery (p=0.10). (b) Lateral movements towards the operative side were not significantly controlled (p=0.54) (Figure 2).

#### 3.2.2. With a traction table

The changes in pelvic alignment in the anteroposterior direction during THA with the use of a traction table are shown in Figure 3. With the traction table, the pelvis tilted forward by 0.2±1.1° on average during anterior capsular release, 0.8±1.5° during femoral head resection, 0.3±1.2° during cup placement, and 0.6±0.9° during reduction. The change averaged 0.2±1.1° from AR registration to anterior capsular release, 0.5±1.4° from anterior capsular release to femoral head resection, -0.4±1.4° from femoral head resection to cup placement, and 0.2±1.0° from cup placement to reduction. The anteroposterior directional changes were not statistically significant, indicating that pelvic movements were more stable intraoperatively with the traction table (p=0.10).

The changes in lateral pelvic alignment at each stage of THA using a traction table are also depicted in Figure 3. The pelvis tilted towards the operative side by 0.9±1.0° on average during anterior capsular release, 1.6±1.1° during femoral head resection, 2.0±1.2° during cup placement, and 2.3±0.8° during reduction. The pelvis consistently tilted towards the operative side throughout the procedure. The change averaged 0.9±1.0° from AR registration to anterior capsular release, 0.2±1.5° from anterior capsular release to femoral head resection, 0.4±1.5° from femoral head resection to cup placement, and 0.4±1.2° from cup placement to reduction. No significant difference in the amount of lateral movement was observed, showing a gradual tilt towards the operative side throughout the procedure (p=0.54) (Figure 3).

### 3.3. Standard table vs traction table

Table 2 presents a comparison of the anteroposterior movements at various surgical stages using the standard and traction tables, whereas Table 3 shows a comparison of the lateral movements (operative and non-operative sides) at various surgical stages using the standard and traction tables. In patients who underwent THA using the standard table compared to those using the traction table, the traction table significantly stabilized the movements in the anteroposterior direction at all surgical stages (p<0.05), except during anterior capsular release. When comparing the lateral pelvic movements, the pelvic tilt to the operative or non-operative direction at any surgical stage did not significantly differ between the standard and traction table groups. This result indicated that although the traction table effectively stabilized the anteroposterior pelvic movements, it did not significantly affect the lateral movements during THA.

**Table 2.** Comparison of anteroposterior movements at various surgical stages using the standard table and traction table.

| Surgical stages | Standard table (°) | Traction table (°) | *p*-value |
| --- | --- | --- | --- |
| Anterior capsular release | 0.9±1.1 | 0.2±1.3 | 0.053 |
| Femoral head resection | 3.4±0.8 | 0.8±1.5 | <0.05 |
| Cup placement | 2.3±0.8 | 0.3±1.2 | <0.05 |
| Reduction | 2.0±0.8 | 0.6±0.9 | <0.05 |

**Table 3.** Comparison of lateral movements (operative and non-operative sides) at various surgical stages using the standard table and traction table.

| Surgical stages | Standard table (°) | Traction table (°) | *p*-value |
| --- | --- | --- | --- |
| Anterior capsular release | 1.0±0.8 | 0.9±1.0 | 0.63 |
| Femoral head resection | 1.9±1.3 | 1.6±1.1 | 0.40 |
| Cup placement | 2.0±1.3 | 2.0±1.2 | 0.93 |
| Reduction | 2.8±0.7 | 2.3±0.8 | 0.17 |

## 4. Discussion

This study demonstrated that using a traction table significantly stabilized anteroposterior pelvic movements during THA, contributing to more precise acetabular cup alignment.

### 4.1. Pelvic tilt and cup alignment in THA

Pelvic tilt has been suggested to influence the alignment of the acetabular component in THA [13]. Intraoperative pelvic alignment varies owing to manipulation during the procedure and patient factors such as BMI [14,15]. An unstable pelvis intraoperatively complicates precise cup placement, leading to potential malalignment, increased risk of dislocations, and premature wear of the implant [3]. Our study highlights the importance of real-time monitoring and stabilization of pelvic movement to ensure accurate cup placement.

### 4.2. Utilization of the traction table in THA

The use of a traction table in THA allows for controlled femoral rotation and facilitates exposure of the femoral bone [16]. Previous studies have reported high rates of greater trochanteric fractures with the anterior approach without traction tables [17] and suggested the need for additional incisions to expose the acetabulum and femur in > 3,000 cases using the Heuter approach on a standard table [16]. In contrast, when a traction table was used, no additional skin incisions were necessary when positioning the acetabular component [16]. Thus, the use of a traction table in THA simplifies the approach to the femur, potentially reduces skin incisions, and minimizes the risk of muscle damage caused by forced traction.

### 4.3. Effectiveness of the traction table in THA

Our study revealed that the use of the traction table stabilized the anteroposterior pelvic movements intraoperatively. For every degree of anterior pelvic tilt, there is an associated increase of 0.21° and 0.73° in the abduction and anteversion angles of the cup [18]. Therefore, stabilizing pelvic motion with a traction table can enhance postoperative cup alignment.

### 4.4. Consideration of anterior pelvic tilt during osteotomy

The anterior pelvic tilt observed during osteotomy is thought to be a phenomenon caused by a combination of multiple factors, including changes in the pelvic support structure and muscle tone, as well as the effects of surgical manipulation and the operating table. Understanding these factors and responding appropriately intraoperatively will enable more accurate acetabular cup placement and contribute to improved surgical outcomes.

### 4.5. Limitations of the traction table in THA

Our findings also indicated that the traction table did not stabilize lateral pelvic movement during THA. Each degree of lateral tilt towards the operative side increased the abduction and anteversion angles by 0.28° and 0.63°, respectively [18]. Because the traction table does not control lateral motion, there is a risk of significant malalignment in the abduction and anteversion angles of the cup, leading to suboptimal placement.

### 4.6. Pelvic alignment in THA

Although anteroposterior pelvic alignment can be controlled, lateral movements cannot be stabilized during THA. Improper cup alignment is primarily caused by intraoperative pelvic movement [19]. Therefore, it is crucial to be aware of pelvic movement intraoperatively and take precautions to minimize pelvic tilting.

Our study elucidates how the pelvis moves intraoperatively with and without the use of the traction table, and highlights the differences in pelvic dynamics between the two. Portable navigation systems can track pelvic motion in real time. Without such systems, understanding the dynamic changes in pelvic alignment at each surgical stage would enable more accurate cup placement.

### 4.7. Limitations and future research

This study was limited to the use of a specific traction table, and the results for other types of traction tables or surgical approaches remain unknown. The accuracy and limitations of the portable navigation system used to track pelvic movements in real time have not been thoroughly validated. An adequate analysis on how variables such as patients’ anatomical characteristics and BMI influence pelvic movements and cup alignment intraoperatively has not been conducted. Although data on short-term surgical outcomes were collected, this study did not include information on long-term outcomes. Thus, the relationship between acetabular cup alignment, long-term functionality, and durability remains unclear. This study utilized a retrospective design; prospective studies or randomized controlled trials are necessary. Future studies should address these limitations and involve a more diverse patient population and a range of surgical approaches. In addition, the development of new technologies and surgical instruments to control pelvic movements more precisely is critical.

### 4.8. Conclusions

This study examined pelvic motion during THA and the impact on precise acetabular cup alignment. Specifically, it focused on the differences in pelvic movements when using a standard table versus a traction table and how these affected the surgical outcomes. The results confirmed that the use of a traction table clearly controlled the anteroposterior pelvic movements, contributing to accurate acetabular cup alignment. This underscores the importance of pelvic stability intraoperatively, which is essential for the success of THA, particularly for proper acetabular cup placement. However, lateral pelvic movements are not controlled by the traction table, leaving room for further research and technological development.

## Data Availability Statement

The datasets used and/or analyzed during the current study are available from the corresponding author on reasonable request.

## Acknowledgements

None.

